# Altered hippocampal functional connectivity patterns in patients with cognitive impairments following ischaemic stroke: a resting-state fMRI study

**DOI:** 10.1101/2020.10.26.20219782

**Authors:** JeYoung Jung, Rosanna Laverick, Kurdow Nader, Martin Wilson, Dorothee P. Auer, Pia Rotshtein, Akram A. Hosseini

**Affiliations:** School of Psychology, University of Nottingham, UK; School of Psychology, University of Birmingham, UK; University Hospital Birmingham NHS Trust, Birmingham, UK; NIHR Nottingham BRC, University of Nottingham, UK; Division of Clinical Neuroscience, University of Nottingham, UK; Department of Neurology, Nottingham University Hospitals NHS Trust, Queen’s Medical Centre, Nottingham, UK

**Keywords:** Ischemic stroke, cognitive impairment, hippocampus, resting-state functional magnetic resonance imaging, functional connectivity

## Abstract

**Background:** Ischemic stroke with cognitive impairment is a considerable risk factor for developing dementia. Identifying imaging markers of cognitive impairment following ischemic stroke will help to develop prevention strategies against post-stroke dementia.

**Methods:** Here, we investigated the hippocampal functional connectivity (FC) pattern following ischemic stroke, using resting-state fMRI (rs-fMRI). Thirty-three cognitively impaired patients after ischemic stroke and sixteen age-matched controls with no known history of neurological disorder, were recruited for the study. Importantly, no patient had a direct ischaemic insult to hippocampus on examination of brain imaging. Seven subfields of hippocampus were used as a seed region for FC analyses.

**Results:** Across all hippocampal subfields, FC with the inferior parietal lobe in patients was reduced as compared with healthy controls. This decreased FC included both supramarginal gyrus and angular gyrus. The FC of hippocampal subfields with cerebellum was increased. Importantly, the degree of the altered FC between hippocampal subfields and IPL was associated with their impaired memory function.

**Conclusion:** Our results demonstrated that decreased hippocampal-IPL connectivity was associated with cognitive impairment in patients with ischemic stroke. These findings provide novel insights into the role of hippocampus in cognitive impairment following ischemic stroke.

## 1. Introduction

Stroke is a leading cause of death and disability word-widely and ischemic stroke accounts for around 71% of all strokes (Gorelick et al., 2011). Despite intensive treatment, and rehabilitation, more than half of the patients with ischemic stroke cannot return to work due to the remaining physical or cognitive dysfunction (Campbell et al., 2019). Patients with ischemic stroke are at increased risk of developing vascular cognitive impairment ranging from mild cognitive impairment (MCI) to vascular dementia (Rockwood et al., 1999). In particular, stroke patients with cognitive impairment have an increased risk of developing vascular or other types of dementia at 5 years (Wentzel et al., 2001). Although studies demonstrated a significant link between ischemic stroke with cognitive impairment, and vascular dementia (Rockwood et al., 2000), its underlying neural mechanism remain unclear. It is important to identify markers for developing cognitive impairment in ischemic stroke, which might be beneficial to prevent from progression to vascular dementia.

Hippocampal integrity is a well-established diagnostic marker for dementia and Alzheimer’s disease (AD) (den Heijer et al., 2010; Elcombe et al., 2014; La Joie et al., 2014). Changes to the structure and function of the hippocampus is considered predictive of progression from MCI to AD (Barnes et al., 2009). As the hippocampus plays a major role in learning and memory, hippocampal changes (e.g., atrophy or decreased activation) are a key factor in the process of age-related memory and cognitive impairment (Fotuhi et al., 2012; O’Brien et al., 2010). Furthermore, these hippocampal alterations have been associated with cognitive decline or dementia after stroke (Blum et al., 2012; Gemmell et al., 2012; Yang et al., 2014). Werden et al (2017) reported hippocampal volume loss 6 weeks after ischemic strokes as compared to healthy controls. Hippocampus is composed of cytoarchitectonically different subfields such as cornu ammonis (CA1-CA3), subiculum, and dentate gyrus (DG) (Amunts et al., 2005). Studies have reported that each subfields plays a specific role within the hippocampal circuitry (Berron et al., 2016; Hodgetts et al., 2017; Neunuebel and Knierim, 2014). Especially, in the early stage of AD, neuron loss are more prominent in the CA1 and subiculum (Rossler et al., 2002). The CA3 and DG was associated with auditory immediate recall, while CA1 was related to auditory delayed recall in temporal lobe epilepsy patients with hippocampal sclerosis (Mueller et al., 2012). However, the subfields of hippocampal functional changes after an ischemic stroke and their association with cognitive impairment remain less clear.

Resting-state functional magnetic resonance imaging (rs-fMRI) measures the temporal correlation of spontaneous low-fluctuations (typically < 0.1Hz) in the blood oxygenation level-dependent (BOLD) signals among different brain areas, providing a measure of temporal coherence between brain regions at rest – functional connectivity (FC). Recent rs-fMRI studies of stroke patients have demonstrated disruptions in the FC in the lesioned and peri-lesioned parenchyma, followed by connectivity-based reorganization, and subsequent functional recovery (Baldassarre et al., 2016; Kroll et al., 2017; Ovadia-Caro et al., 2014). The interhemispheric FC reduced after acute ischemia, which correlated with patients’ impaired cognitive performance (Carter et al., 2010; Puig et al., 2018). Specifically, the motor network FC was decreased in patients with ischemic stroke (Golestani et al., 2013) and the FC improved alongside with motor function recovery (Wang et al., 2010; Zheng et al., 2016). Sun et al (2011) investigated the FC of default mode network (DMN) after ischemic stroke in patients with cognitive impairment. Their analyses showed decreased FC of the precuneus with multiple brain regions such as frontal, temporal and subcortical areas; as well as increased FC with the middle/inferior temporal gyrus. Another study examined the hippocampal FC and its change related to cognitive therapy (computer-assisted cognitive rehabilitation) after ischemic stroke (within 6months) (Yang et al., 2014). The patients in their study had abnormal hippocampal FC with insula, cerebellum, prefrontal and temporal cortex, compared with controls. After cognitive therapy, the hippocampal FC with prefrontal cortex and precuneus increased in the patient group, and was associated with improved memory function. A study of patients with transient ischemic attack revealed decreased amplitude of low frequency fluctuation – spontaneous brain activity - in middle temporal gyrus, compared to controls (Lv et al., 2019). These studies showed that even in those with transient ischemic attack, the event induces FC alterations in the various brain regions. However, there is little known about the impact of FC following an ischemic stroke.

In the current study, we employed rs-fMRI to explore the pattern of hippocampal FC after an ischemic stroke in patients with post-stroke cognitive impairment. To provide detailed hippocampal FC, we used the cytoarchitectonic probabilistic maps of hippocampus, which divides it into 7 subfields (CA1, CA2, CA3, DG, EC, HATA, and Subiculum) (Amunts et al., 2005; Eickhoff et al., 2005). We hypothesized that there could be altered patterns of hippocampal FC in patients after ischemic stroke as compared with controls. The degree of changes in the hippocampal FC would be associated with their impaired memory function.

## 2. Materials and Methods

### 2.1 Participants

We used data from the Hippocampal Pathology in Post-Stroke Cognitive Impairment study (HiPPS-CI) (a total of 71 stroke patients and 20 age-matched controls) acquired from two West-Midlands hospitals (Queen Elizabeth Birmingham and Sandwell General Hospital) between July 2015 and January 2019.

Thirty-three patients (6 females, mean age 62.5 ± 13.4 years) were included in this study following inclusion criteria. Our inclusion criteria for the study were (a) recent (less than three months) clinically diagnosed ischemic stroke, (b) age >18 to <90 years, (c) able and willing to provide informed consent and (d) cognitive impairment (Montreal cognitive assessment MoCA <=26/30. Stroke patients were excluded from the study if they (a) had contraindication to have MRI e.g. metal foreign body (pacemaker, aneurysm clip, possibility of metal fragments in the eye, etc), (b) unfit or unable to tolerate MRI e.g. unable to lie flat due to backache or severe kyphosis, shortness of breath, (c) Severe disabling stroke (m-Rankin Scale > 4) (Fish, 2011), (d) known pre-stroke dementia or cognitive impairment as confirmed by family members or medical documents, (e) if their lesion directly affected the hippocampus region. Stroke patients were recruited within their hospital admission. At this stage informed consent was taken, and clinical and demographic information recorded. They were invited to attend Birmingham University Imaging Centre to take part in a cognitive assessment, and MRI at 3T within three months of stroke.

Sixteen age-matched controls with no previous history of neurological disorder (8 females, mean age = 61.2 ± 9.5 years) were recruited during the same period, they were recruited as relatives of stroke patients or from the local community.

The study was approved by the UK Health Research Authority and West Midlands Black Country Research Ethics Committee (15/WM/0209). All participants provided signed written informed consent prior to the study.

### 2.2 Clinical and cognitive evaluation

Clinical profiles were obtained from the clinical records of the patients. The control group provided self-report information of their health. For each participant we computed a vascular risk score based on the Framingham stroke risk profile (FSRP). FSRP is an estimate of the individual’s stroke risk in the next 10 years, which represents a level of vascular health (Wolf et al., 1991). FSRP includes the following risk factors: age, systolic blood pressure (taken at admission to hospital), antihypertensive medication, diabetes, cigarette smoking, history of cardiovascular disease, and atrial fibrillation. A higher vascular risk score indicated worse prognosis for further stroke incidence, and lower overall vascular health. Cognition was assessed using Montreal Cognitive Assessment, MoCA (Nasreddine et al., 2005) and Birmingham Cognitive Screen, BCoS (Bickerton et al., 2015; Humphreys, 2012; Pan et al., 2015). The BCoS assessed the participants’ cognitive profiles across five cognitive domains: (a) attention and executive function, (b) language, (c) memory, (d) number, and (e) praxis from 22 tasks. It should be noted that some participants could not complete the sub-questions of each task. A test of verbal fluency (Lezak, 1976) was also included in the cognitive battery, as it is widely used in assessment of neurological patients (Henry and Crawford, 2004; Herbert et al., 2014).

In addition, each participant’s functional independence was assessed by using the Barthel index (Mahoney and Barthel, 1965). The hospital anxiety and depression scale (HADS) (Zigmond and Snaith, 1983) was utilised to assess the participants’ mood. Cognitive and mental health screening were performed during their hospital admission as part of clinical care after stroke for Barthel, or by Clinical Research Nurses or the first assessment for MoCA and HADS.

### 2.3 Statistical analysis

Differences in demographics and clinical scales between the stroke patients and controls were analysed using a t-test for continuous variables (age, years of education, FSRP, MoCA, Barthel, HADS, and verbal fluency) and χ^2^ test for gender.

The BCoS scores were analysed employing analysis of covariance (ANCOVA) with covariates including the age, gender, and years of education.

All statistical analyses were performed using IBM SPSS Statistics for Windows, version 24.0 (IBM Corporation, Armonk, NY, USA).

### 2.4 Image acquisition

A 3T Philips Achieve scanner was used to acquire imaging data hosted in Birmingham University Imaging centre. During the rsfMRI, participants were asked to keep their eyes closed, lie as still as possible and to stay awake (6mins, the total volume: 180). Anatomical images were acquired the MPRAGE sequence (TR = 8.4ms, TE = 3.8ms, matrix = 288 × 288, resolution = 1 × 1 × 1 mm^3^) covering the whole head. rsfMRI were obtained using a single-shot echo planer imaging (EPI) sequence (TR = 2000ms, TE = 35ms, matrix = 80 x 80, number of slices = 32, resolution = 3 × 3 × 3mm^3^). The scanning protocol included additional image sequences (e.g. T2, FLAIR, MRS, DTI) which will be reported elsewhere. The rsfMRI was the 7^th^ sequence and was collected at about 30 minutes into the scanning.

### 2.5 Lesion analysis

Identification and quantification of the ischemic lesions was performed manually by two raters using MRIcroGL (https://www.nitrc.org/projects/mricrogl). It was guided by clinical imaging reporting by NK. The number of voxels of the lesion site ROI was multiplied by the voxels of the scan (lesion voxels x 0.56 x 0.56 x1); the lesion was measured in mm^3^. The lesion volume is presented as percentage of intracranial volume obtained from Computation Anatomy Toolbox (CAT12, http://www.neuro.uni-jena.de/cat/). The results are summarised in Table S1 (see, Supplementary Information). It should be noted that, no patient had a direct ischaemic insult to hippocampus.

### 2.6 Image pre-processing

Image pre-processing was performed using statistical parametric mapping (SPM 12) software (Wellcome Trust Centre for Neuroimaging) and the Data Processing Assistant for Resting State fMRI (DPARSF Advanced Edition, version 2.3) toolbox (Yan et al., 2016).

SPM 12 was used for slice timing correction, realignment, and coregistration to the individual’s structural image. Participants with > 2 mm translation or 1 degree of rotation were excluded from the analysis. To reduce the effect of head movements, we used four methods: censoring, global signal regression, 24-motion parameter regression, and scrubbing of high motion time points. Within the DPARSF, nuisance covariates were regressed out and the images were normalized using DARTEL (Ashburner, 2007) and smoothed with and 5mm full-width half maximum Gaussian kernel. The results were filtered at 0.01–0.08 Hz (Satterthwaite et al., 2013).

Nuisance covariates included 24 motion parameters calculated from the six original motion parameters using Volterra expansion (Friston et al., 1996). Time points with a z-score > 2.5 from the mean global power or > 1 mm translation were identified as outliers using the ARtifact detection Tools software package (ART;www.nitrc.org/projects/artifact_detect). Each of these was entered as a covariate. White matter, cerebrospinal fluid, and global tissue signal were covaried out and linear detrending was performed to reduce motion-related artifacts (Anderson et al., 2011; Power et al., 2014; Weissenbacher et al., 2009). Four participants (3 patients and 1 control) were excluded due to having motion > 2 mm of translation, 1° of rotation, or < 6 min of data remaining after scrubbing high motion time points.

### 2.7 Functional connectivity analysis

Seed-based functional connectivity analyses were performed using the DPARSF. For the sub-fields of hippocampus, we used the template from the Anatomy toolbox’s cytoarchitectonic probabilistic maps (Fig. 1) (Amunts et al., 2005; Eickhoff et al., 2005). The sub-fields of bilateral hippocampus were included in this analysis as a seed region (CA1, CA2, CA3, DG, EC, HATA, and Subiculum). To generate functional connectivity (FC) maps, bivariate correlations were calculated between each seed and the whole brain voxels. The correlation coefficient map was converted into z map by Fisher’s r-to-z transform to improve the normality (Rosner, 2006). The resultant FC maps were generated for the stroke patients and the control groups using one-sample t-test and compared between the groups using t-tests in the SPM 12, accounting for age, gender, and the years of education. The statistical threshold was set at p _FDR-corrected_ < 0.05, Ks > 50 at a cluster level and p < 0.001 at a voxel level. In addition, we conducted a conjunction analysis across the sub-regions of hippocampus between the groups. The statistical threshold was set at p _FDR-corrected_< 0.05, Ks > 50 at a cluster level and p _FWE-corrected_ < 0.05 at a voxel level.

**Figure 1.**
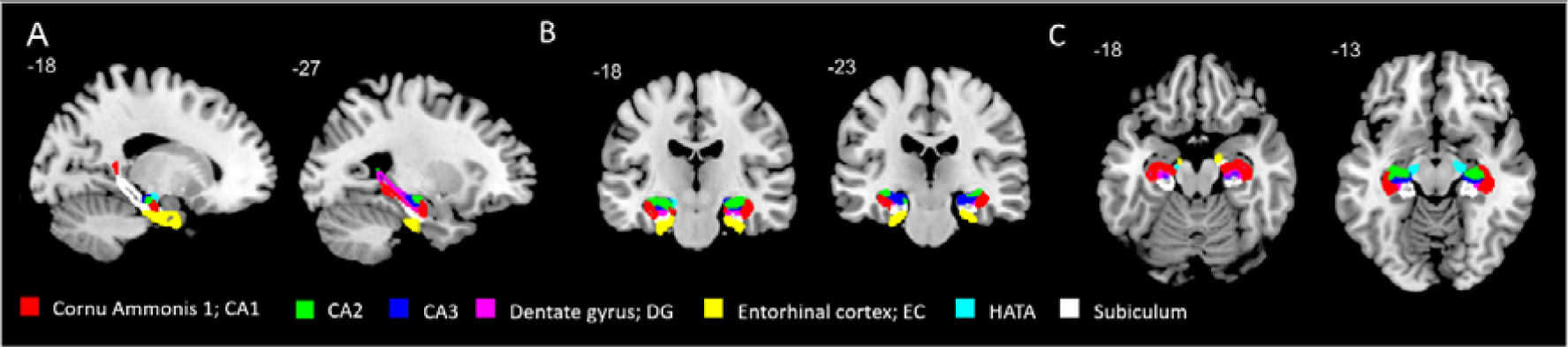
The subfields of hippocampus.

In order to explore the relationship between the altered FC and patients’ impaired cognitive functions, the Functional Connectivity (CONN) Toolbox (http://web.mit.edu/swg/software.htm) was used. Regions of interest (ROIs) were defined based on the seed-based functional connectivity results (conjunction analysis results, see Fig. 2). Five ROIs were used from the automated anatomical labelling (AAL) template (Rolls et al., 2020) including supramarginal gyrus (anterior and posterior), angular gyrus, superior parietal lobe, and cerebellum (Fig. 3). Pre-processed images were entered to the toolbox with the covariates (age, gender, and the years of education). The sub-regions of hippocampus were also included as ROIs. ROI-to-ROI analysis was performed between the hippocampal sub-regions and ROIs by calculating the bivariate correlation between the each pair of ROIs. Then, correlation analysis was conducted to link the FC changes between ROIs and individual’s impaired memory function, accounting for age, gender, and the years of education (p _FDR-corrected_ < 0.05).

**Figure 2.**
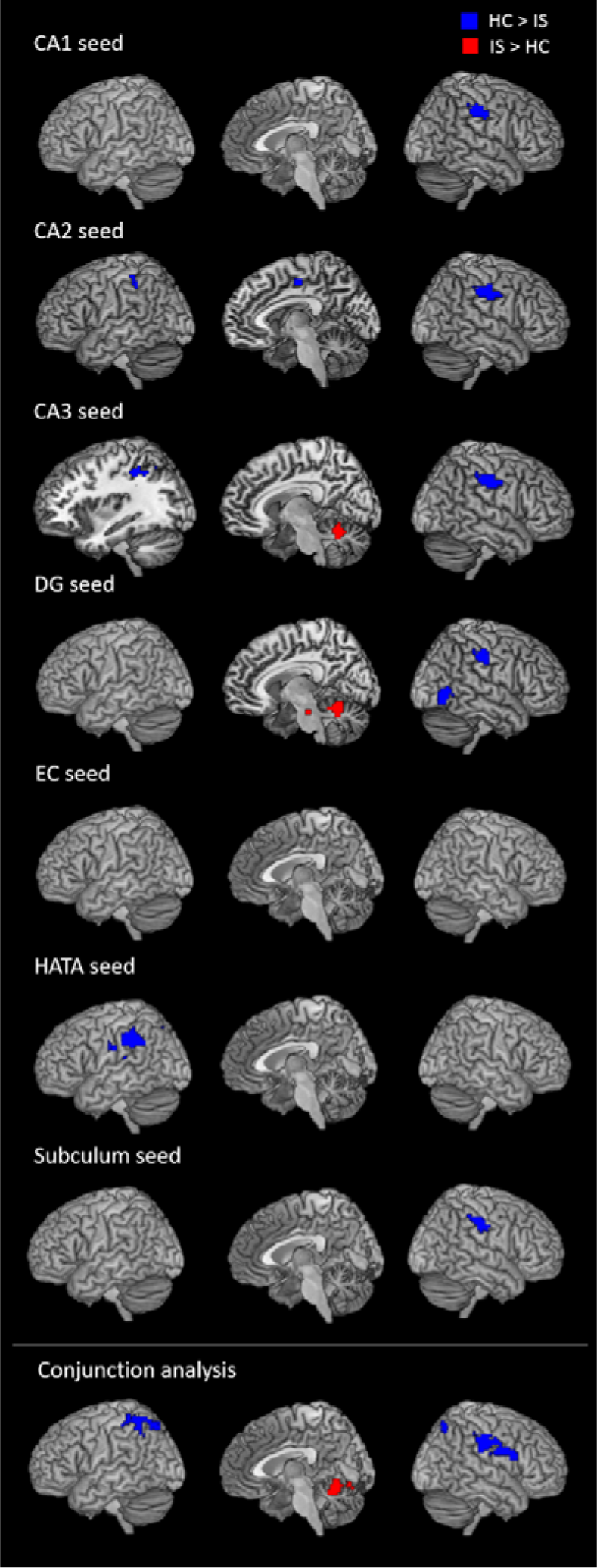
The results of functional connectivity pattern of hippocampal subfields.

**Figure 3.**
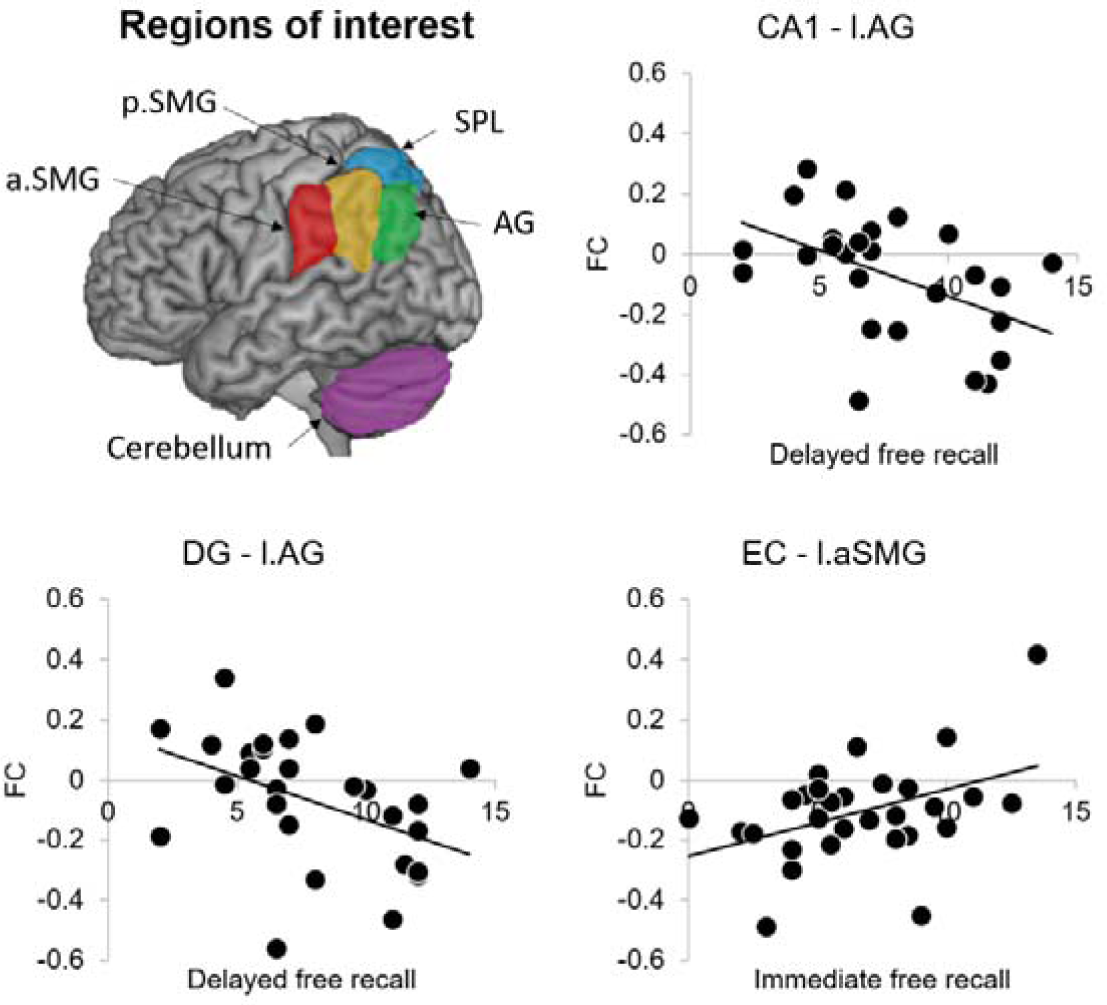
The relationship between the functional connectivity of hippocampal subfields and memory function in the patients. aSMG: anterior supramarginal gyrus, pSMG: posterior supramarginal gyrus, AG: angular gyrus, SPL: superior parietal lobe

## 3. Results

### 3.1 Demographics, clinical, and cognitive scores

Table 1 summarises the results of demographics, clinical and cognitive scores. There was no age difference between the patients and control groups (p > 0.7). However, there was a significant difference in the years of education (t = -2.40, p = 0.021) and gender (χ^2^ = 6.35, p = 0.017) between the two groups.

**Table 1.** Demographic information, clinical and cognitive scores.

|  | HC<br>(n = 16) | IS<br>(n = 33) | Statistics | p value |
| --- | --- | --- | --- | --- |
| Demographics: mean (SD) |  |  |  |  |
| Gender, male/female | 5/9 | 25/7 | $\chi^2 = 6.35$ | **0.017\*** |
| Age | 60.38 (9.33)<br>(n = 13) | 62.27 (13.02)<br>(n = 33) | t = 0.48 | 0.637 |
| Education, year | 14.61 (3.09)<br>(n = 13) | 12.36 (2.67)<br>(n = 28) | t = -2.40 | **0.021\*** |
| Clinical scales: mean (SD) |  |  |  |  |
| FSRP | 12.61 (12.26)<br>(n = 13) | 25.63 (14.13)<br>(n = 32) | t = 2.90 | **0.006\*\*** |
| HADS Anxiety | 2.50 (1.76)<br>(n = 6) | 6.14 (4.54)<br>(n = 29) | t = 1.91 | 0.065 |
| HADS Depression | 1.00 (0.89)<br>(n = 6) | 5.45 (3.84)<br>(n = 29) | t = 5.55 | **< 0.001\*\*\*** |
| Barthel | 20.00 (0)<br>(n = 6) | 20.58 (11.79)<br>(n = 29) | t = 0.12 | 0.905 |
| MoCA | 26.57 (2.30)<br>(n = 7) | 21.36 (4.89)<br>(n = 33) | t = -2.73 | **0.009\*\*** |
| BCoS scores: mean (SD) |  |  | F |  |
| Attention |  |  |  |  |
| Apple cancellation | 48.2 (1.8) | 47.8 (3.6) | 0.31 | 0.580 |
| Left visual bilateral | 8.0(0) | 7.8 (0.6) | 0.17 | 0.686 |
| Right visual bilateral | 8.0(0) | 7.7 (1.5) | 0.00 | 0.975 |
| Left tactile bilateral | 8.0(0) | 7.8 (0.4) | 1.06 | 0.314 |
| Right tactile bilateral | 8.0(0) | 7.9 (1.8) | 0.05 | 0.824 |
| Rule finding and switching | 11.7 (3.0) | 10.4 (3.9) | 0.28 | 0.600 |
| Auditory attention accuracy | 53.1 (1.3) | 51.3 (4.9) | 1.28 | 0.270 |
| Memory |  |  |  |  |
| Personal information | 8 (0) | 7.7 (1.5) | 0.04 | 0.840 |
| Time and space | 5.9 (0.3) | 5.7 (1.2) | 0.30 | 0.590 |
| Immediate free recall | 9.2 (1.8) | 6.6 (3.1) | 8.14 | **0.008\*\*** |
| Immediate recognition | 13.6 (1.8) | 12.8 (1.5) | 0.51 | 0.605 |
| Delayed free recall | 10.6 (1.6) | 7.9 (3.3) | 5.08 | **0.033\*** |
| Delayed recognition | 13.5 (1.5) | 12.5 (1.9) | 0.77 | 0.388 |
| Task recognition | 9.9 (0.2) | 9.4 (0.8) | 6.24 | **0.019\*** |
| Figure copy | 46.0 (2.1) | 43.3 (3.0) | 5.95 | **0.022\*** |
| Language |  |  |  |  |
| Picture naming | 13.5 (0.9) | 11.8 (3.1) | 4.54 | **0.042\*** |
| Sentence construction | 7.9 (0.5) | 7.4 (1.6) | 0.42 | 0.520 |
| Sentence reading (accuracy) | 39.9 (7.2) | 39.9 (7.0) | 0.23 | 0.933 |
| Sentence reading (second) | 13.7 (3.6) | 23.0 (11.7) | 8.36 | **0.007\*\*** |
| Nonword reading (accuracy) | 4.9 (2.1) | 4.8 (1.5) | 0.48 | 0.857 |
| Nonword reading (second) | 5.8 (1.1) | 13.5 (8.3) | 14.40 | **0.001\*\*\*** |
| Word writing | 4.7 (0.8) | 3.5 (1.4) | 12.71 | **0.002\*\*** |
| Number |  |  |  |  |
| Number reading | 9 (0) | 8.5 (0.7) | 3.36 | 0.078 |
| Number writing | 4.9 (0.4) | 4.8 (0.8) | 0.05 | 0.820 |
| Calculation | 3.4 (1.1) | 3.3 (0.9) | 1.79 | 0.190 |
| Praxis |  |  |  |  |
| Multiple object use | 11.9 (0.3) | 11.8 (0.6) | 0.77 | 0.440 |
| Gesture prouction | 11.8 (0.6) | 11.2 (1.4) | 1.40 | 0.150 |
| Gesture recognition | 5.8 (0.6) | 5.7 (0.7) | 0.28 | 0.770 |

IS: ischemic stroke patient, HC: healthy control, FSRP: Framingham stroke risk profile, HADS: hospital anxiety and depression scale, MoCA: Montreal cognitive assessment, BCoS: Birmingham cognitive screen. \* $p < 0.05$ , \*\* $p < 0.01$ , \*\*\* $p < 0.001$

The FSRP, MoCA and HADS scores were significantly different between two groups (FSRP: t = 2.90, p = 0.006; MoCA: t = -2.73, p = 0.009; HADS Anxiety: t = 1.91, p = 0.065; HADS Anxiety: t = 5.84, p < 0.001). There was no difference in the Barthel index between the groups (p > 0.9).

The BCoS scores revealed that the stroke patients had impairments in their memory and language function compared to the controls. In the memory function, the stroke group scored lower in the immediate free recall (F_4, 29_ = 8.14, p = 0.008), delayed free recall (F_4, 28_ = 5.08, p = 0.033), and task recognition (F_4, 25_ = 6.24, p = 0.019) relative to the control group. In the language domain, the patients with stroke showed worse performance in picture naming (F_4, 29_ = 4.54, p = 0.042), sentence (F_4, 29_ = 8.36, p = 0.007) and nonword reading (F_4, 28_ = 14.40, p < 0.001) compared with control. The patients also scored lower in Complex Figure Copy compared to the controls (F_4, 26_ = 5.95, p = 0.022). There was no significant difference in the other domains (Fs< 0.001, ps> 0.97).

### 3.2 Functional connectivity results

Seed-based functional connectivity analysis is displayed in the Fig. 2 and Table 2. The overall results revealed decreased hippocampal FC with the supramarginal gyrus (SMG) and angular gyrus (AG), and increased FC with the cerebellum in the stroke group compared to the control group. Additionally, the patients showed FC reduction between CA2-supplementary motor area and between DG-right inferior occipital gyrus. The EC FC did not show any difference between the groups.

**Table 2.**
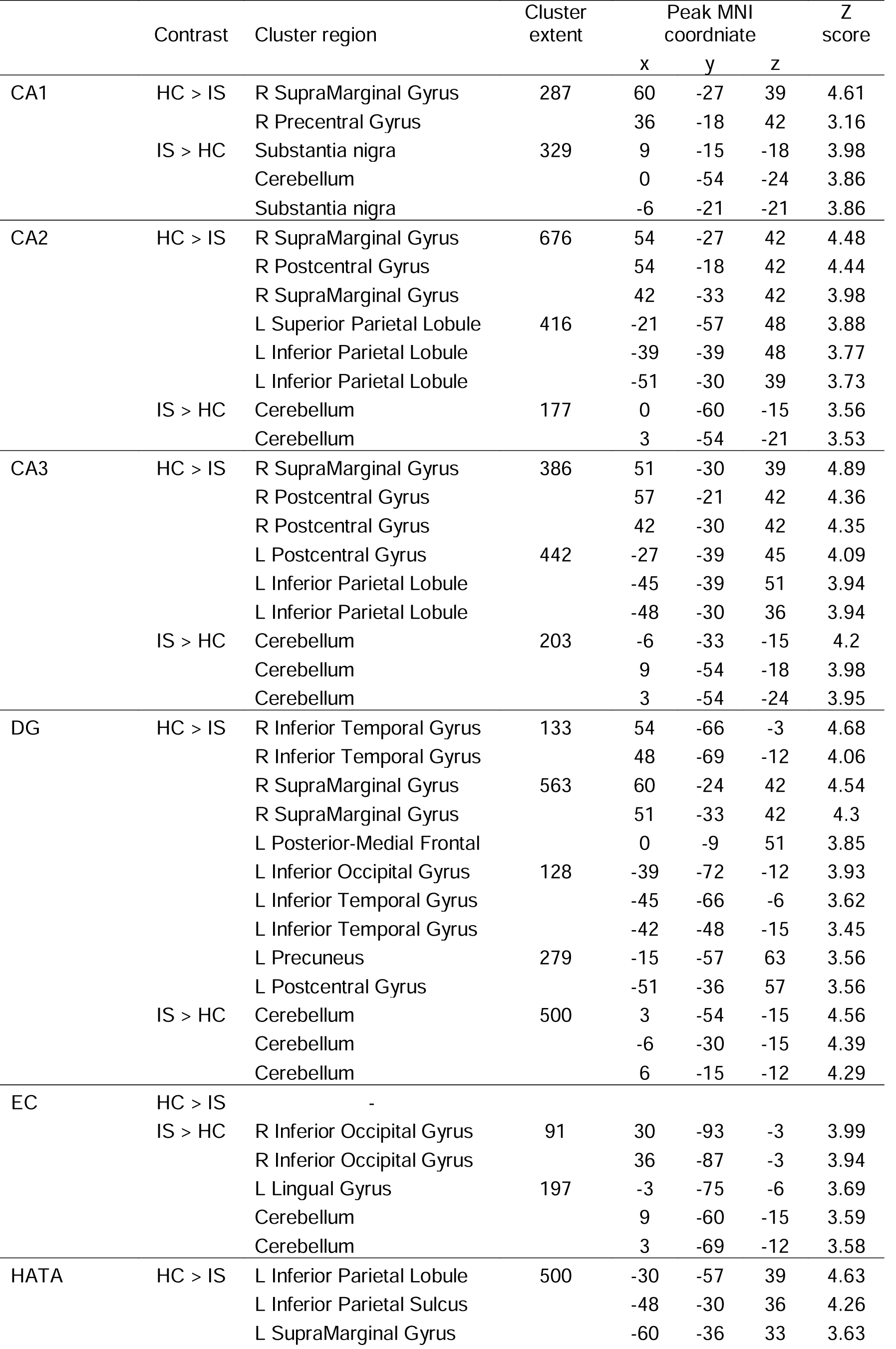
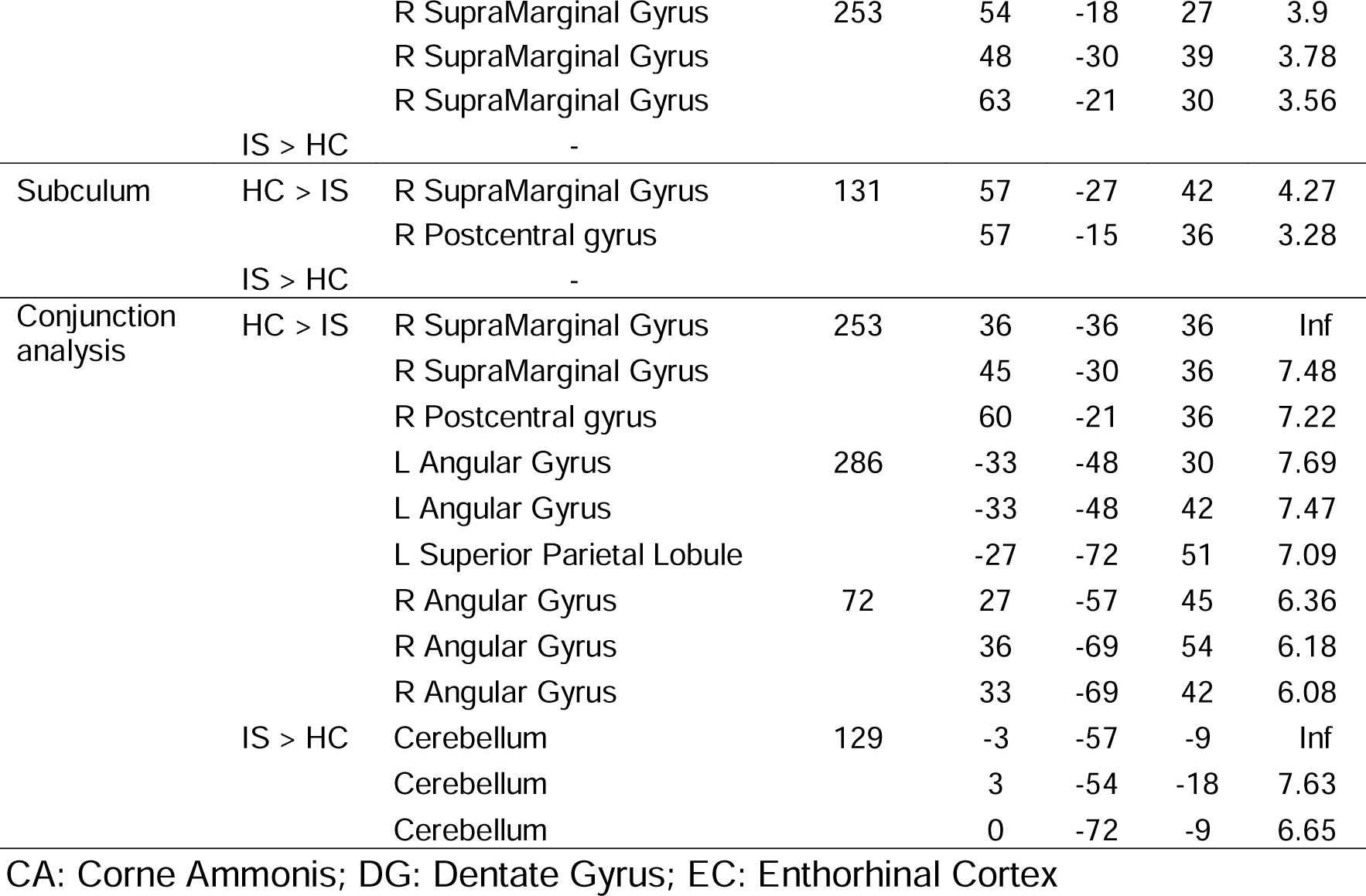
The results of seed-based FC analysis.

In order to find the overlapping areas across the FC of the hippocampal subfields, a conjunction analysis was conducted. The results demonstrated that relative to the controls, FC between the hippocampus-SMG and hippocampus-AG decreased in the stroke group, while FC between the hippocampus and cerebellum increased.

### 3.3 The relationship between the altered FC and impaired cognitive functions in patients

We explored the relationship between the altered FC found in the stroke group and their cognitive scores (Fig. 3). We extracted the FC between the hippocampal sub-regions and ROIs (anterior SMG: aSMG, posterior SMG: pSMG, AG, and cerebellum) and correlated with the impaired memory function (immediate free recall, and delayed free recall, see Table 1), accounting for age, gender, and the years of education.

The analysis revealed that the delayed free recall score in the stroke patients was negatively correlated with the CA1-left AG FC (r = -0.51, p _FDR-corrected_ = 0.025) and DG-left AG FC (r = -0.45, p _FDR-corrected_ = 0.06). The more decreased FC in the CA1-left AG and DG-left AG in the patients, the better score in the delayed free recall was observed. The stroke group showed a significant correlation between the EC-left aSMG FC and immediate free recall score (r = 0.42, p _FDR-corrected_ = 0.049). The patients with stronger FC in the EC-left aSMG performed better in the immediate free recall. The control group showed no significant correlations (CA1-left AG & delayed free recall: r = -0.49, p _FDR-corrected_ = 0.07, DG-left AG & delayed free recall: r = -0.46, p _FDR-corrected_ = 1, EC-left aSMG & immediate free recall: r = 0.36, p _FDR-corrected_ = 1) (Fig. S1).

## 4. Discussion

The current study examined the altered FC of the hippocampal subfields in patients with post-stroke cognitive impairment. The altered FC of hippocampal subfields was mainly located in the inferior parietal lobe (IPL) and cerebellum. Furthermore, the degree of the functional connectivity changes between the hippocampal subfields and IPL was associated with post-stroke cognitive impairment involving memory. These findings provide novel insights into our understanding of cognitively impaired patients following an ischemic stroke (post-stroke cognitive impairment), as compared with age-matched healthy controls.

The novel finding of the current study is decreased FC between the hippocampal subfields and inferior parietal lobe (IPL) regions (supramarginal gyrus: SMG and angular gyrus: AG) after ischemic stroke, compared with control group. The SMG is the anterior part of the IPL and associated with visual attention (Chambers et al., 2004; Stevens et al., 2005), working memory (Sommer et al., 2006; Uncapher and Wagner, 2009), motion perception (Martinez-Trujillo et al., 2007), and, phonological (Humphreys and Ralph, 2015; Oberhuber et al., 2016) and semantic processing (Price, 2010; Raposo et al., 2006). Stroke patients with left SMG lesions demonstrated impairments in their verbal memory function (Beeson et al., 1993; Caplan et al., 1995). A study with amnestic MCI patients investigated the hippocampal FC compared to controls and reported altered hippocampal FC in frontal, temporal, and parietal lobe such as the SMG (Bai et al., 2009). Similarly, we found that post-stroke memory impairment was associated with functional connectivity alterations between the hippocampal subfields and SMG. These hippocampal FC changes might be the early sign of functional abnormalities in the episodic memory system (Bai et al., 2009). Furthermore, the patients in our cohort with more decoupling in the hippocampus-SMG FC performed worse in immediate free recall when compared with controls. Our findings suggest that the dysfunctional relationship between the hippocampus and SMG may contribute to post-stroke memory impairment and could be a potential neural marker during subacute ischaemic stroke.

AG is the posterior part of the IPL and a cross-modal hub (Grayson et al., 2014; Seghier, 2013) supported by its widespread structural and functional connection with various brain regions (Seghier et al., 2010). Thus, the function of the AG is diverse ranging from attention, spatial cognition, language, semantic processing, and social cognition to episodic memory (Binder et al., 2009; Humphreys and Ralph, 2015; Price et al., 2015; Seghier, 2013; Vincent et al., 2006). In addition, functional connection of the AG reveals that the AG is a key region of the default mode network (DMN) as well as a connector hub across the DMN, salience, executive control, and attention networks (Humphreys et al., 2015; Igelstrom and Graziano, 2017; Xu et al., 2016). Newhart et al (2012) studied patients with acute ischemia and showed the working memory deficit in the patients with the AG lesion. Transcranial magnetic stimulation (TMS) studies also demonstrated the causal role of the left AG in episodic memory by delivering inhibitory protocols over the left AG (e.g., temporarily disrupting ongoing processing at the target brain region) (Bonnici et al., 2018; Thakral et al., 2017). Another study of post-stroke cognitive impairment (Tuladhar et al., 2013) reported a decreased connectivity within the DMN including the hippocampus compared to controls. Snaphaan et al (2009) examined the working memory function in stroke patients and reported that hippocampal activation was reduced during 2-back working memory task. These findings highlight the role of the AG and the interaction between the hippocampus and AG in episodic memory function. Similarly, we found that patients with ischaemic stroke had lower FC between the hippocampal subfields and AG. Further, the degree of memory impairment after stroke correlated with the level of the FC alterations. Taken together, our findings suggest the dysfunctional connectivity of hippocampus underlying memory dysfunction after ischemic stroke.

Emerging evidence from functional neuroimaging, neurophysiology and computational modelling supports the importance of interaction between the hippocampus, medial temporal cortex, IPL, precunues, and prefrontal cortex for memory function (Clower et al., 2001; Cooper and Ritchey, 2019; Geib et al., 2017; King et al., 2015; Simons and Spiers, 2003). Any lesion or disconnection in this network may result in memory dysfunction, which can be found in stroke and dementia patients (Bai et al., 2009; Beeson et al., 1993; Caplan et al., 1995; den Heijer et al., 2010; Elcombe et al., 2014; La Joie et al., 2014; Newhart et al., 2012; Snaphaan et al., 2009; Tuladhar et al., 2013). Thus, patients with ischemic stroke and memory dysfunction could have abnormalities in this system. Our study highlights the decreased hippocampal-IPL connectivity in post-stroke cognitive impairment.

We found an increased FC between the hippocampal subfields and cerebellum. The cerebellum has been related to motor control, cognition, and emotion (Glickstein, 2007; Strick et al., 2009). Furthermore, the cerebellum is involved in storing formed memory, and known to show an increased activation during memory tasks (Groussard et al., 2010; Kuper et al., 2016). Cerebellar-hippocampal interaction is associated with various cognitive functions such as spatial and temporal processing (For a reivew, see Yu and Krook-Magnuson, 2015) and a few studies have reported increased cerebellar-hippocampal FC during a finger tapping task (Onuki et al., 2015), eye blink conditioning (Singer, 1999), and spatial navigation (Rochefort et al., 2011). These studies indicate the importance of cerebellum and the cerebellar-hippocampal connection for cognitive tasks. Previously, Yang et al (Yang et al., 2014) showed increased hippocampal FC with insula, medial frontal gyrus, superior temporal gyrus, and cerebellum in patients with unilateral infarction of the basal ganglia. Alternatively, the enhanced hippocampal-cerebellar FC in our study might be driven from a potential compensatory mechanism after ischaemic stroke.

Several limitations in this study need to be considered. First, the patients differed from the controls in sex and years of education. Although we included them as covariates in all of our analysis, future studies are required with better matched and higher number of healthy controls. Second, the patients had higher depression score than the controls. The post-stroke depression has been reported to alter FC in the DMN, cognitive control network and affective network (Lassalle-Lagadec et al., 2012; Zhang et al., 2018). To confirm our findings, we re-analysed our data with HADS depression scores as an additional covariate. The results replicated our initial findings – the decreased hippocampal-IPL connectivity and the increased hippocampal-cerebellum connectivity in the patients. Thus, our results were not affected by the depression in the patient group.

Clinically, our finding could contribute to the identification of patients with post-stroke cognitive impairment due to the altered hippocampal FC patterns, before emerging irreversible hippocampal atrophy. It might have significant implications in diagnostics because many of stroke patients with cognitive impairment are at increased risk of delayed hippocampal atrophy and developing post-stroke dementia (Rockwood et al., 2000; Wentzel et al., 2001), which is three times higher than the risk of a recurrent stroke (Yokota et al., 2004).

## Supporting information

manuscript

## Data Availability

All the data in the manuscript are available and held in the University of Birmingham (Centre for Human Brain Health).

## Author contributions

PR, DPA and AAH conceptualised the study and designed the research; RL recruited participants and obtained the data; KN and MW ensured MRI quality control. J.J analysed the data and performed the statistical analysis; J.J, RL, PR, and AAH wrote the manuscript. All co-authors edited the manuscript.

## Acknowledgement

We would like to acknowledge study co-investigators not listed as co-authors, the research nurses at Sandwell General Hospital and Queen Elizabeth Hospital Birmingham, and the Birmingham University Imaging Centre (now the Centre for Human Brain Health) for supporting the study. Additionally, Rachel Evans and students; Emily Todd and Alicia Northall for the support in data collection.

