## Supplementary material for "Altered hippocampal functional connectivity patterns in patients with cognitive impairments following ischaemic stroke: a resting-state fMRI study": manuscript

**Supplementary information**

Supplementary Figure 1

Supplementary Tables 1

**Supplementary Figure 1**

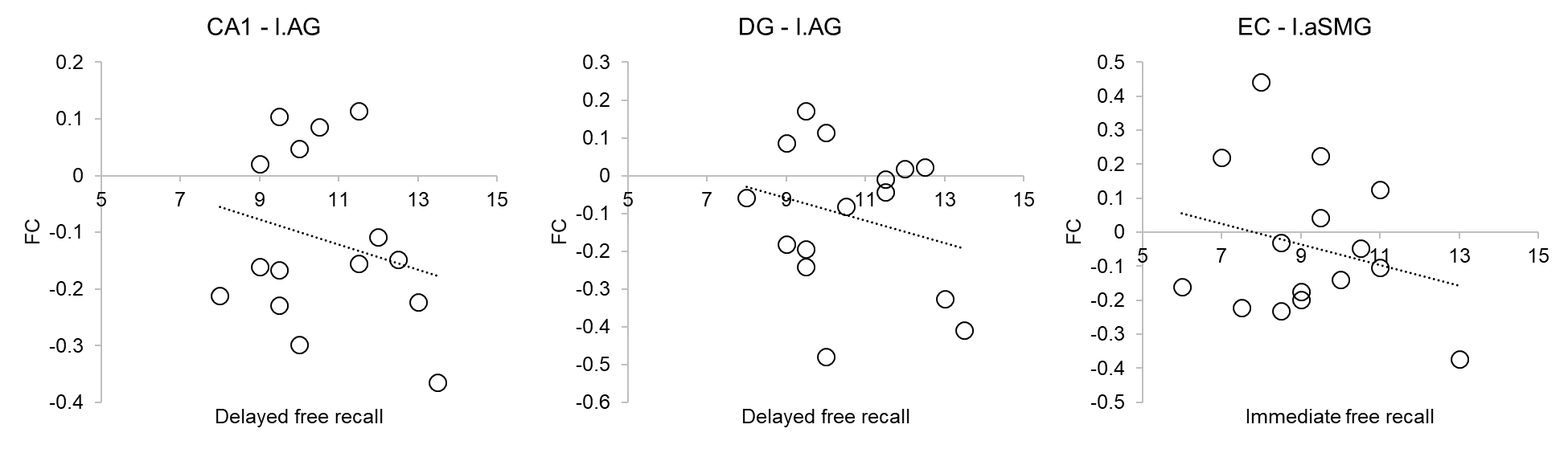

**Figure S1.** Relationship between the hippocampal FC and memory function in HCs

**Supplementary Table 1**

| ID | Lesion Side | Lesion Location | Lesion Volume | Specification |
| --- | --- | --- | --- | --- |
| IS01 | Left | Cortical | 0.05 | / |
| IS02 | Left | Cortical | 0.12 | Left occipital Lobe |
| IS03 | Left | Cortical | 2.37 | Left frontal lobe including cortex, small area of posterior partial lobe |
| IS04 | Left | Cortical | 0.01 | Left frontal Lobe |
| IS05 | Left | Cortical | 0 | / |
| IS06 | Bilateral | Cortical | 0.2 | Left frontal lobe, occipital and temporal lobes. Right posterior parietal lobe |
| IS07 | Right | Subcortical | 0.03 | Infarct in right lentiform nucleus |
| IS08 | Left | Cortical | 0.06 | Left posterior frontal lobe/ parietal region |
| IS09 | Left | Subcortical | 0.02 | Left lacunar infarct |
| IS10 | Left | Subcortical | 0.15 | Basal Ganglia intracranial haemorrhage |
| IS11 | Right | Cortical | 0.07 | Right parietal-occipital lobe |
| IS12 | Bilateral | Subcortical | 0 | Small foci infarcts in both hemispheres, left thalamus |
| IS13 | Right | Subcortical | 0.02 | Lacunar infarct in posterior of right ventricle. |
| IS14 | Left | Subcortical | 0.03 | Pontine haemorrhage |
| IS15 | Left | Subcortical | 0.12 | Left cerebellum and adjacent left pons |
| IS16 | Left | Subcortical | 0 | / |
| IS17 | Right | Cortical | 0.11 | Right parietal lobe, basal ganglia and insular cortex |
| IS18 | Left | Subcortical | 0.01 | Left inferior pons |
| IS19 | Left | Cortical | 0.04 | Left posterior MCA territory of parietal lobe |
| IS20 | Right | Cortical | 0.69 | Large right frontal lobe infarct, insular cortex and part of parietal lobe |
| IS21 | Right | Cortical | 0.01 | Occipital lobe with tiny foci posteriorly in right parietal lobe |
| IS22 | Left | Subcortical | 0.03 | inferomedial portion of left cerebella hemisphere, PICA territory |
| IS23 | Left |  | 0 | / |
| IS24 | Right | Cortical | 0.04 | medial parietal (retrosplenial cortex) |
| IS25 | Right | Cortical | 1.48 | frontal (facial droop) |
| IS26 | Bilateral | Subcortical | 0.01 | Bilateral thalamic infarction |
| IS27 | Right | Subcortical | 0.03 | Striatal region (caudate, anterior lentiform nucleus) |
| IS28 | Right | Cortical | 0.49 | Right superior cerebral artery |
| IS29 | Left | Cortical | 0.1 | Posterior frontal infarct |
| IS30 | Left | Cortical | 0.04 | / |
| IS31 | Left | Cortical | 0.04 | posterior temporal gyrus and parietal |
| IS32 | Left | Subcortical | 0.09 | Cerebellum |

**Table S1.** Summary of lesion identification and quantification in IS patients
